# Sleep duration, physical activity and cardiometabolic multimorbidity: Findings from the English Longitudinal Study of Ageing

**DOI:** 10.1101/2025.01.30.25321441

**Authors:** Setor K. Kunutsor, Jari A. Laukkanen

## Abstract

**Background:** U-shaped relationships have been demonstrated between sleep duration and several cardiometabolic conditions. However, the association between sleep duration and cardiometabolic multimorbidity (CMM) and the interrelationship with physical activity has not been previously investigated. We aimed to assess the prospective associations of sleep habits (duration and quality) with CMM and the interplay with physical activity.

**Methods and Results:** We included 3,428 participants (mean age 63 years at baseline, 44.8% male) free of hypertension, coronary heart disease, diabetes, and stroke at wave 4 from the English Longitudinal Study of Ageing. Sleep habits were assessed by self-reports. Cardiometabolic multimorbidity was defined as the presence of at least two multiple long-term conditions (hypertension, cardiovascular disease, diabetes, and stroke) at wave 10. Odds ratios (ORs) with 95% confidence intervals (CIs) were estimated using logistic regression models adjusted for established cardiometabolic risk factors including physical activity. At 15 years follow-up, 206 participants developed CMM. There was an approximate U-shaped trend between sleep duration and the risk of CMM. Compared to participants with sleep duration of 7-8 hrs/day, the multivariable OR (95% CI) for CMM was 1.39 (1.03-1.90) for sleep duration ≤6 hrs/day and 1.05 (0.55-2.00) for sleep duration ≥ 9 hrs/day. The odds of CMM modestly decreased for each additional hour increase in sleep duration in participants with sleep duration ≤6 hrs/day (OR, 0.78, 95% CI: 0.59-1.02). Sleep quality or physical activity was not associated with CMM.

**Conclusions:** Short sleep duration is associated with an increased risk of CMM, which is independent of established cardiometabolic risk factors including physical activity. Importantly, each additional hour of sleep among those with short sleep durations appeared to mitigate CMM risk, underscoring the potential benefits of optimizing sleep duration.

---

Cardiometabolic conditions such as type 2 diabetes (T2D), hypertension, and cardiovascular disease (CVD) represent significant global health burdens, contributing substantially to morbidity, mortality, and high healthcare costs.^1^ The co-occurrence of at least two of these conditions, termed cardiometabolic multimorbidity (CMM), is becoming increasingly prevalent due to the aging population and the growing prevalence of lifestyle-related risk factors.^2^ Cardiometabolic multimorbidity not only exacerbates individual morbidity and mortality risks but also poses considerable challenges to healthcare systems worldwide.^3^ Patients with CMM are at higher risk of complications, more frequent hospitalizations, and poorer quality of life compared to those with a single condition,^4^ further underscoring the urgency of addressing its risk factors. The development of CMM is strongly influenced by advancing age, obesity, and modifiable lifestyle factors such as physical inactivity, smoking, excessive alcohol consumption, and unhealthy dietary habits.^5^

Increasing attention has also been directed toward poor sleep habits as a novel and modifiable risk factor for cardiometabolic health.^6,7^ Specifically, both short (<7 hours) and prolonged (>8 hours) sleep durations have been linked to an increased risk of type 2 diabetes (T2D) and cardiovascular disease (CVD), while 7–8 hours of sleep is associated with the lowest risk, following a U-shaped relationship.^8–10^ Sleep quality, including difficulty in initiating or maintaining sleep, has similarly been linked to heightened risks of cardiometabolic conditions.^10,11^ Recognizing the importance of sleep, the American Heart Association (AHA) recently updated its cardiovascular health metrics, now termed Life’s Essential 8 (LE8), to include sleep health as a key determinant.^12^ These developments emphasize the potential impact of improving sleep habits on reducing cardiometabolic risks, though evidence regarding the relationship between sleep patterns and CMM remains limited. Despite growing evidence of the relationship between sleep habits and individual cardiometabolic conditions, the association between sleep duration and CMM is less clear. Given the multifactorial etiology of CMM, the interplay between poor sleep and other protective factors, such as physical activity, warrants investigation. Physical activity is a well-established protective factor against chronic diseases, including hypertension, T2D, and CVD, ^13–16^ and higher physical activity levels are associated with lower risks of CMM.^17,18^ Emerging evidence suggests that physical activity may mitigate the adverse effects of common cardiovascular risk factors, including poor sleep patterns.^19^ This study hypothesizes that short sleep duration is associated with an increased risk of CMM and that the association is influenced by physical activity levels, with higher levels of physical activity attenuating or mitigating the increased risk of CMM associated with short sleep duration. Using data from the English Longitudinal Study of Ageing, this study aims to investigate the association between sleep duration and the risk of CMM, with a focus on the interplay between sleep and physical activity. In a secondary analysis, we explored the relationship between sleep quality and CMM.

## Materials and Methods Study

### Design and Population

This research was carried out adhering to the STROBE (Strengthening the Reporting of Observational Studies in Epidemiology) guidelines, which are standards for reporting epidemiological observational studies (**Table S1**).^20^ Study participants in this analysis were drawn from English Longitudinal Study of Ageing (ELSA), an ongoing, population-based, prospective cohort study comprising a nationally representative sample of the English population living in households.^21^ Study participants were recruited from the Health Surveys for England, a selection of annual, population-based surveys carried out in 1998, 1999, and 2001.^21^ At inception in 2002–2003, the ELSA core sample comprised 11,391 nationally representative males and females aged ≥50 years who were living in private residential households in England. The ELSA sample has been followed up every 2 years (referred to as wave) since the inception of the cohort.^21^ For the purposes of the present analyses, data collected during wave 4 (2008–2009) were used as the baseline, given that this was the first wave that collected data on sleep habits. The follow-up in the current study ended at wave 10 (2021-2023). For this analysis, we excluded participants with prevalent hypertension, coronary heart disease (CHD), diabetes, or stroke. Complete data on sleep habits (duration and quality), physical activity, covariates, and CMM status were available for 3428 men and women. Participants provided full informed written consent to participate in the study and ethical approval was obtained from the London Multicentre Research Ethics Committee.

### Assessment of Exposures, Covariates and Outcome

Sleep duration for each participant was collected through the baseline questionnaire with the question: “How many hours of sleep you typically had on an average weeknight”. Sleep duration was categorized into three groups: ≤6 hours (short sleep duration), 7-8 hours (optimal sleep duration), and ≥9 hours (long sleep duration), for consistency with previous studies.^10^ Sleep quality was evaluated by four questions: (1) How often do you feel it was hard to fall asleep during the last month; (2) How often do you wake up several times at night during the last month; (3) How often do you feel tired and worn out at wake up in the morning during the last month; and (4) Overall sleep quality rating.

Overall sleep quality was classified into four categories: very good, good, fairly bad, and very bad. Due to the small sample sizes in the “fairly bad” and “very bad” groups, these were combined into a single “bad” category. Data on age, sex, physical activity, alcohol consumption, smoking status, and self-reported medical conditions were collected using standardized questionnaires in ELSA. Physical activity was initially classified into four categories—sedentary, low, moderate, and high—based on the frequency of participation in vigorous, moderate, and mild sports or activities.^8,21^ For this analysis, it was reclassified into three categories—low, moderate, and high—due to the small sample sizes in the sedentary and low activity groups. Heights, weights, and blood samples were measured during physical examinations conducted at the Mobile Examination Centre by nurses using standardized protocols. Body mass index (BMI) was calculated as weight (kg) divided by height squared (m²). The diagnoses of hypertension, CHD, diabetes, and stroke were based on participants’ self-reported medical histories. Cardiometabolic multimorbidity was defined as the presence of at least two of these long-term conditions (hypertension, CVD, diabetes, and stroke) at wave 10.

### Statistical Analysis

Baseline characteristics were presented as either means (with standard deviation, SD) or medians (with interquartile range, IQR) for continuous variables, and as numbers (with percentages) for categorical variables. Group comparisons of continuous and categorical variables were performed using independent-sample *t*-tests and chi-square tests, respectively. To explore a potential nonlinear dose-response relationship between sleep duration and CMM risk, we constructed a multivariable restricted cubic spline (RCS) with knots at the 5th, 35th, 65th, and 95th percentiles of the distribution of sleep duration as recommended by Harrell.^22^ Binary logistic regression analyses were used to estimate odds ratios (ORs) and 95% confidence intervals (CIs) for the associations of sleep habits (duration and quality) and physical activity with CMM status at follow-up. For the associations of sleep duration categories with the risk of CMM, the category 7-8 hrs/day was used as the reference comparison, as previous studies of sleep and cardiometabolic diseases have shown that this is the optimal sleep duration range associated with the lowest risk of adverse outcomes.^8,9,23^ Covariate adjustments were made using three progressive models: (Model 1) age and sex; (Model 2) Model 1 plus smoking status, systolic blood pressure (SBP), total cholesterol, high-density lipoprotein cholesterol (HDL-C), and BMI; and (Model 3) Model 2 plus physical activity. These confounders were selected based on the following criteria: their roles as established risk factors for cardiometabolic diseases, documented associations with cardiometabolic diseases in the ELSA study,^2,8^ and their potential to confound the observed relationships between exposures and outcomes.^24^ All statistical analyses were performed using Stata version MP 18 (Stata Corp, College Station, Texas).

## Results

**Table 1** presents the baseline characteristics of the study’s participants overall and by CMM status at study follow-up. The mean (SD) age of the 3428 study participants at baseline was 63 (9) years and males constituted 44.8% of the cohort. The mean (SD) sleep duration was 7.0 (1.2) hrs/day. Participants who developed CMM had shorter sleep duration, more likely to be smokers, had higher levels of SBP, total cholesterol, and BMI, and had lower levels of HDL-C.

**Table 1.** Characteristics of participants overall and by cardiometabolic multimorbidity status.

| Characteristic | Overall<br>(N=3428) | No CMM<br>(N=3222) | Yes CMM<br>(N=206) | p-value |
| --- | --- | --- | --- | --- |
|  | Mean (SD) or n (%) | Mean (SD) or n (%) | Mean (SD) or n (%) |  |
| Sleep duration, hrs/day | 7.0 (1.2) | 7.0 (1.2) | 6.7 (1.4) | 0.009 |
| Sleep Quality |  |  |  | 0.18 |
| Bad | 688 (20.1%) | 639 (19.8%) | 49 (23.8%) |  |
| Good | 1884 (55.0%) | 1769 (54.9%) | 115 (55.8%) |  |
| Very good | 856 (25.0%) | 814 (25.3%) | 42 (20.4%) |  |
| Age, yrs | 63 (9) | 64 (9) | 63 (7) | 0.15 |
| Sex |  |  |  | 0.16 |
| Male | 1536 (44.8%) | 1434 (44.5%) | 102 (49.5%) |  |
| Female | 1892 (55.2%) | 1788 (55.5%) | 104 (50.5%) |  |
| Current smoker |  |  |  | 0.055 |
| No | 2935 (85.6%) | 2768 (85.9%) | 167 (81.1%) |  |
| Yes | 493 (14.4%) | 454 (14.1%) | 39 (18.9%) |  |
| Alcohol categories |  |  |  | 0.83 |
| None | 1162 (33.9%) | 1088 (33.8%) | 74 (35.9%) |  |
| 1-2 times/wk | 859 (25.1%) | 811 (25.2%) | 48 (23.3%) |  |
| 3-4 times/wk | 647 (18.9%) | 611 (19.0%) | 36 (17.5%) |  |
| 5 or more times/wk | 760 (22.2%) | 712 (22.1%) | 48 (23.3%) |  |
| Systolic blood pressure, mmHg | 130 (17) | 130 (17) | 137 (17) | <0.001 |
| Total cholesterol, mmol/l | 5.84 (1.14) | 5.85 (1.13) | 5.60 (1.19) | 0.002 |
| HDL cholesterol, mmol/l | 1.59 (0.42) | 1.60 (0.42) | 1.45 (0.41) | <0.001 |
| Body mass index, kg/m<sup>2</sup> | 27.6 (5.0) | 27.4 (4.9) | 30.0 (5.9) | <0.001 |
| Physical activity |  |  |  | 0.093 |
| Low | 717 (20.9%) | 662 (20.5%) | 55 (26.7%) |  |
| Moderate | 1845 (53.8%) | 1739 (54.0%) | 106 (51.5%) |  |
| High | 866 (25.3%) | 821 (25.5%) | 45 (21.8%) |  |

CMM, cardiometabolic multimorbidity; HDL, high-density lipoprotein; SD, standard deviation

At 15 years follow-up, 206 participants developed CMM. A multivariable RCS showed a non-linear trend (U-shape) in the relationship between sleep duration and the risk of CMM; the risk of CMM was highest in participants with the shortest sleep duration, with risk decreasing with increasing sleep duration up till 7-8 hours (**Figure 1**). Compared to participants with sleep duration of 7-8 hrs/day, the age and sex-adjusted OR (95% CI) for CMM was 1.50 (1.12-2.02) in participants with sleep duration ≤6 hrs/day (**Figure 1-Model 1**), which was attenuated to 1.40 (1.03-1.90) on further adjustment for smoking status, SBP, total cholesterol, HDL-C, and BMI (**Figure 1-Model 2**). Further adjustment for physical activity had no effect on the association (**Figure 1-Model 3**). Evidence for the association between sleep duration ≥9 hrs/day and risk of CMM was not significant. The odds of CMM modestly decreased for each additional hour increase in sleep duration in participants with sleep duration ≤6 hrs/day (OR, 0.78, 95% CI: 0.59-1.02, *p*=.073) after adjusting for age, sex, smoking status, SBP, total cholesterol, HDL-C, BMI, and physical activity.

**Figure 1.**
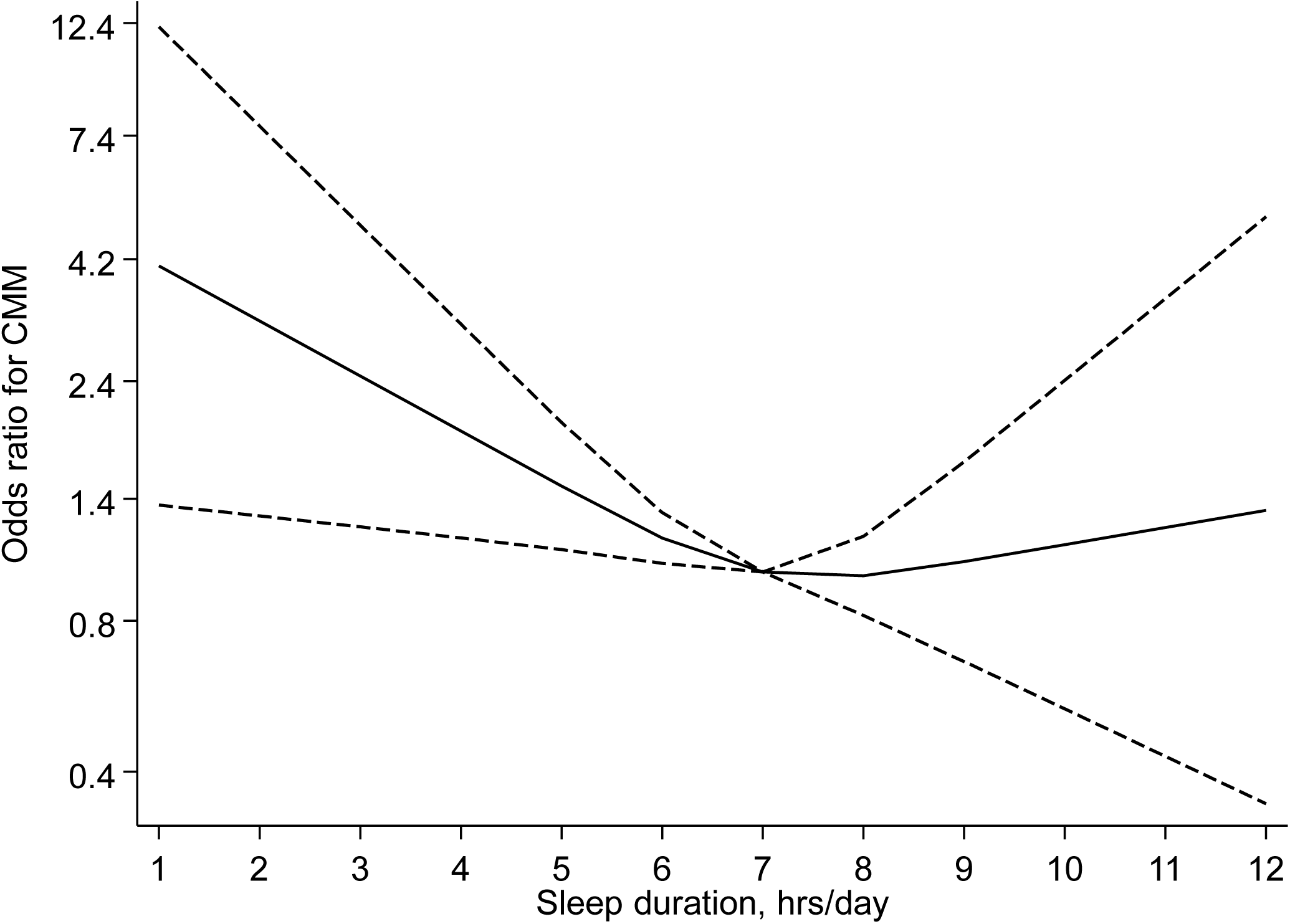
Restricted cubic spline of the odds ratios of cardiometabolic multimorbidity with sleep duration. Dashed lines represent the 95% confidence intervals for the spline model (solid line). Models were adjusted for age, sex, smoking status, systolic blood pressure, total cholesterol, high-density lipoprotein cholesterol, body mass index, and physical activity.

**Figure 2.**
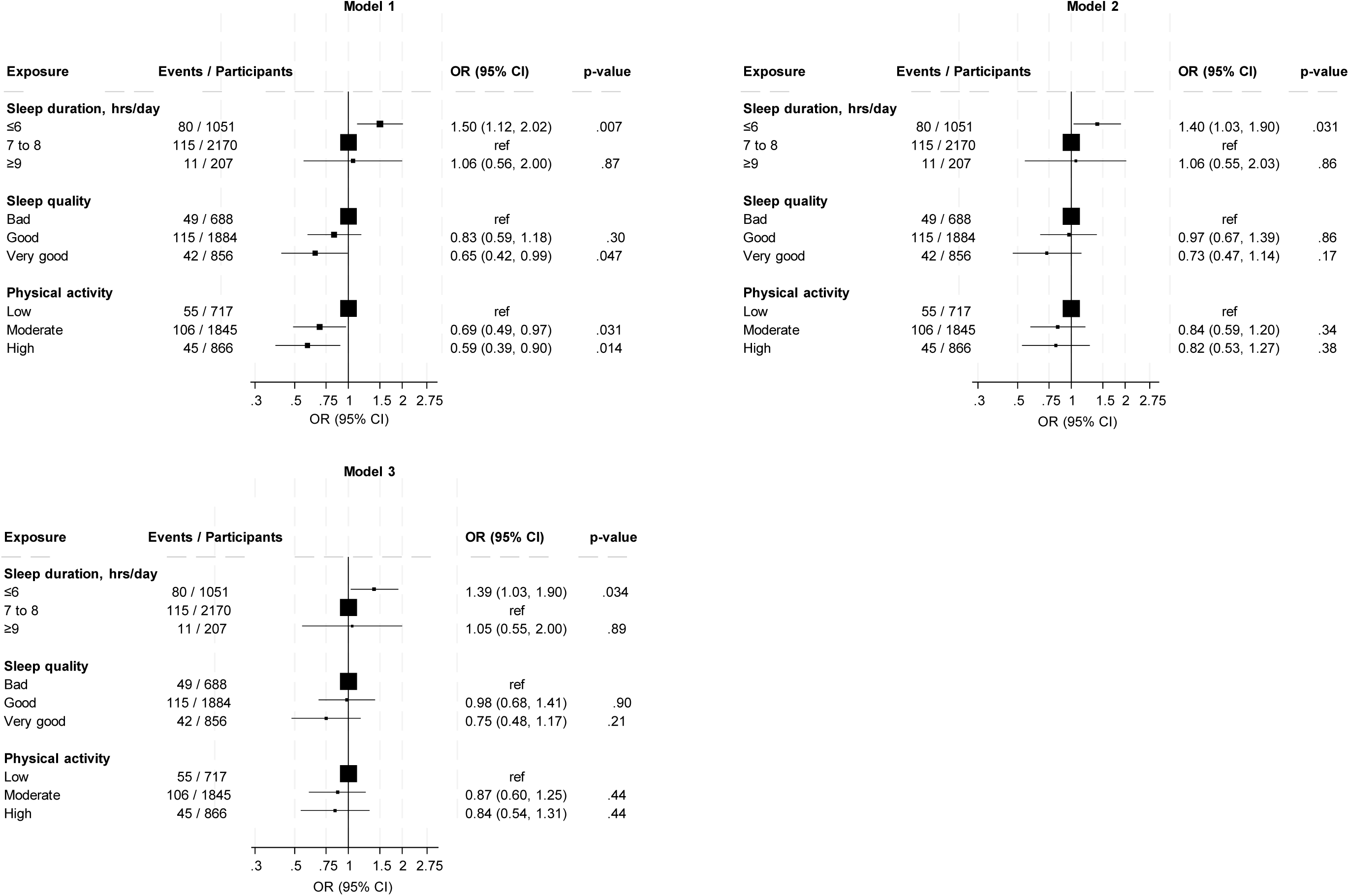
Associations of sleep habits and physical activity with cardiometabolic multimorbidity. CI, confidence interval; OR, odds ratio; ref, reference comparison Model 1: Adjusted for age and sex Model 2: Model 1 plus smoking status, systolic blood pressure, total cholesterol, high-density lipoprotein cholesterol, and body mass index Model 3: Model 2 plus physical activity

Initial evidence of an age and sex-adjusted association between very good vs bad sleep quality and CMM risk (OR, 0.65, 95% CI: 0.42-0.99) (**Figure 1-Model 1**) was attenuated to null on further adjustment for smoking status, SBP, total cholesterol, HDL-C, and BMI (OR, 0.73, 95% CI: 0.47-1.14) (**Figure 1-Model 2**). There was no significant evidence of an association between physical activity and the risk of CMM.

## Discussion

This study investigated the associations between sleep habits and the risk of CMM, as well as the interplay with physical activity, using data from a nationally representative sample of the English population living in households. A dose-response analysis revealed an approximate U-shaped relationship between sleep duration and the risk of CMM, with the highest risk observed in participants with the shortest sleep durations, which declined progressively until stabilizing at 7–8 hours of sleep per day. Short sleep duration (≤6 hours/day) was significantly associated with an increased risk of CMM, independent of established cardiometabolic and lifestyle risk factors, including physical activity. The association between long sleep duration (≥9 hours/day) and CMM risk was not significant. Importantly, in participants with short sleep durations, each additional hour of sleep appeared to mitigate the risk of CMM. Neither sleep quality nor physical activity was associated with CMM risk.

This study is the first to examine the associations between sleep habits and the risk of CMM, making direct comparisons with previous research challenging. However, prior studies have explored the relationship between sleep habits—both duration and quality—and individual cardiometabolic endpoints such as hypertension, T2D, and CVD.^6,10,25^ Consistent with our findings, U-shaped relationships between sleep duration and these endpoints have been frequently documented.^8–10^ Interestingly, the risk gradient from our spline analysis suggests that short sleep duration is associated with a higher risk of CMM than long sleep duration. This contrasts with a recent dose-response meta-analysis of 74 studies that demonstrated a J-shaped relationship between sleep duration and CVD risk, where longer sleep duration was associated with significantly higher mortality and CVD risk than shorter sleep duration.^10^ These discrepancies may reflect differences in the outcomes studied, as CMM comprises multiple cardiometabolic conditions rather than a single endpoint. Furthermore, the inability to identify a significant association between long sleep duration and CMM risk in our study could be due to limited statistical power, given the small sample size and low event rate in the long sleep category (207 participants, 11 events). While we did not observe a significant protective effect of sleep quality on CMM risk, as reported in some previous studies, the direction of the effect estimate hints at a possible protective relationship.

Though the exact mechanisms that underline the association between sleep and adverse cardiometabolic outcomes are not fully understood, some pathways have been postulated. Short sleep duration is associated with a high inflammatory burden, characterized by elevated levels of inflammatory cytokines such as tumour necrosis factor-α and interleukin-6, which impair insulin signaling and glucose tolerance.^26,27^ Sleep restriction also decreases leptin and increases ghrelin levels,^28,29^ promoting appetite, caloric intake, obesity, and impaired glycemic control.^30^ Elevated cortisol levels and altered growth hormone metabolism with reduced sleep may further contribute to adverse cardiovascular risk profiles.^31^ Additionally, abnormal sleep durations are often linked with depressive symptoms, low socioeconomic status, unemployment, and low physical activity, which may confound the observed associations with adverse cardiometabolic outcomes.^32^ Abnormal sleep patterns, including short and long sleep durations, can delay or suppress melatonin secretion, exacerbating insulin resistance and promoting cardiometabolic dysfunction.^33^ While we observed no significant association between sleep quality and CMM risk, this aligns with the inconclusive evidence from previous studies,^10,34^ highlighting the need for further research to clarify these complex relationships. The lack of an observed association between physical activity CMM was unexpected but may be due to several reasons: (i) limitations in accurately capturing physical activity levels, which relied on self-reported measures and may not fully reflect habitual activity or intensity; (ii) inadequate statistical power; and (iii) the effects of physical activity on CMM risk could be mediated through its impact on other cardiometabolic risk factors, which were adjusted for in the analysis, potentially attenuating any direct association.

The findings of this study align with the recommendations of the National Sleep Foundation and the Centers for Disease Control and Prevention, which advocate for 7–9 hours of sleep for adults aged 26–64 years and 7–8 hours for those aged 65 years and older.^35,36^ These results also support the American Heart Association’s guidelines, emphasizing the inclusion of sleep behaviors as part of strategies to improve cardiac and overall health.^6^ The U-shaped trend between sleep duration and CMM underscores the importance of optimal sleep duration in mitigating health risks. Notably, in participants with short sleep durations, each additional hour of sleep was associated with a reduced risk of CMM, highlighting the potential benefits of interventions aimed at extending sleep duration within the optimal range. Taking the overall evidence into consideration suggests that improving both the duration and quality of sleep could play a critical role in reducing the burden of CMM in the population. Future research should focus on large-scale longitudinal studies to validate these findings, explore potential mechanisms, and identify effective strategies to improve sleep habits. Investigating the combined effects of sleep behaviors, physical activity, and other lifestyle factors on CMM risk will be essential to inform targeted clinical and public health interventions.

This study has several notable strengths. It is the first to evaluate the associations between sleep habits and CMM, as well as the interplay with physical activity, providing novel insights into this underexplored area. The prospective cohort design, based on a relatively large UK general population sample with long-term follow-up, strengthens the validity of the findings. Additionally, the comprehensive analysis included adjustments for established cardiometabolic risk factors, such as blood-based lipids and lifestyle factors, and explored dose-response relationships, adding depth to the investigation. However, the study has limitations inherent to its observational design, precluding causal inferences. Although adjustments were made for major risk factors, residual confounding due to unmeasured or inadequately measured variables remains a possibility. The generalizability of the findings is limited to middle-aged and older adults in the UK, and sleep habits were assessed using self-reported data, which may be prone to recall and reporting biases. Furthermore, the relatively small sample size in specific subgroups, such as participants with long sleep durations, and the low event rate may have reduced the statistical power to detect significant associations. Future studies with larger, more diverse cohorts and objective sleep measures are needed to confirm these findings and address these limitations.

## Conclusion

The findings highlight a U-shaped trend between sleep duration and CMM risk, with short sleep duration being a significant risk factor independent of established cardiometabolic and lifestyle factors including physical activity. Importantly, each additional hour of sleep among those with short sleep durations appeared to mitigate CMM risk, underscoring the potential benefits of optimizing sleep duration. While no significant associations were observed for long sleep duration, sleep quality or physical activity, the direction of effect estimates suggests areas for further exploration. These results support existing clinical recommendations for optimal sleep duration and emphasize the importance of integrating sleep behaviors into strategies aimed at reducing the burden of CMM.

## DATA AVAILABILITY STATEMENT

Data from the ELSA are available to the public to download from the UK Data Service at https://ukdataservice.ac.uk/.

## ETHICS STATEMENT

English Longitudinal Study of Ageing Wave 4 received ethical approval from the National Hospital for Neurology and Neurosurgery & Institute of Neurology Joint Research Ethics Committee on 12 October 2007 (07/H0716/48), and all participants provided written informed consent. Ethical approvals for the other waves in the ELSA project can be found on the website: https://www.elsa-project.ac.uk/ethical-approval.

## AUTHOR CONTRIBUTIONS

Conceptualization: Setor K. Kunutsor, Jari A. Laukkanen

Formal analysis: Setor K. Kunutsor

Writing – First draft: Setor K. Kunutsor

Investigation: Setor K. Kunutsor, Jari A. Laukkanen

Methodology: Setor K. Kunutsor, Jari A. Laukkanen

Project administration: Setor K. Kunutsor

Writing – review & editing: Setor K. Kunutsor, Jari A. Laukkanen

---

**Table S1.** STROBE 2007 Statement—Checklist of items that should be included in reports of cohort studies.

| Section/Topic | Item # | Recommendation | Reported on page # |
| --- | --- | --- | --- |
| Title and abstract | 1 | (a) Indicate the study’s design with a commonly used term in the title or the abstract | Page 1 |
|  |  | (b) Provide in the abstract an informative and balanced summary of what was done and what was found | Page 2 |
| Introduction |  |  |  |
| Background/rationale | 2 | Explain the scientific background and rationale for the investigation being reported | Page 4 |
| Objectives | 3 | State specific objectives, including any prespecified hypotheses | Page 4 |
| Methods |  |  |  |
| Study design | 4 | Present key elements of study design early in the paper | Study design and population |
| Setting | 5 | Describe the setting, locations, and relevant dates, including periods of recruitment, exposure, follow-up, and data collection | Study design and population |
| Participants | 6 | (a) Give the eligibility criteria, and the sources and methods of selection of participants. Describe methods of follow-up | Study design and population |
|  |  | (b) For matched studies, give matching criteria and number of exposed and unexposed | Not applicable |
| Variables | 7 | Clearly define all outcomes, exposures, predictors, potential confounders, and effect modifiers. Give diagnostic criteria, if applicable | Assessment of exposures, covariates and outcome |
| Data sources/<br>measurement | 8* | For each variable of interest, give sources of data and details of methods of assessment (measurement). Describe comparability of assessment methods if there is more than one group | Assessment of exposures, covariates and outcome |
| Bias | 9 | Describe any efforts to address potential sources of bias | Statistical analyses |
| Study size | 10 | Explain how the study size was arrived at | Statistical analyses |
| Quantitative variables | 11 | Explain how quantitative variables were handled in the analyses. If applicable, describe which groupings were chosen and why | Statistical analyses |
| Statistical methods | 12 | (a) Describe all statistical methods, including those used to control for confounding | Statistical analyses |
|  |  | (b) Describe any methods used to examine subgroups and interactions | Statistical analyses |
|  |  | (c) Explain how missing data were addressed | Not applicable |
|  |  | (d) If applicable, explain how loss to follow-up was addressed | Not applicable |
|  |  | (e) Describe any sensitivity analyses | Statistical analyses |
| **Results** |  |  |  |
| Participants | 13* | (a) Report numbers of individuals at each stage of study—eg numbers potentially eligible, examined for eligibility, confirmed eligible, included in the study, completing follow-up, and analysed | Study design and population |
|  |  | (b) Give reasons for non-participation at each stage | Study design and population |
|  |  | (c) Consider use of a flow diagram | Study design and population |
| Descriptive data | 14* | (a) Give characteristics of study participants (eg demographic, clinical, social) and information on exposures and potential confounders | Results; Table 1 |
|  |  | (b) Indicate number of participants with missing data for each variable of interest |  |
|  |  | (c) Summarise follow-up time (eg, average and total amount) | Results |
| Outcome data | 15* | Report numbers of outcome events or summary measures over time | Results |
| Main results | 16 | (a) Give unadjusted estimates and, if applicable, confounder-adjusted estimates and their precision (eg, 95% confidence interval). Make clear which confounders were adjusted for and why they were included | Results; Figures 1-2 |
|  |  | (b) Report category boundaries when continuous variables were categorized | Results; Table 1; Figure 1-2 |
|  |  | (c) If relevant, consider translating estimates of relative risk into absolute risk for a meaningful time period |  |
| Other analyses | 17 | Report other analyses done—eg analyses of subgroups and interactions, and sensitivity analyses | Results |
| **Discussion** |  |  |  |
| Key results | 18 | Summarise key results with reference to study objectives | Discussion |
| **Limitations** |  |  |  |
| Interpretation | 20 | Give a cautious overall interpretation of results considering objectives, limitations, multiplicity of analyses, results from similar studies, and other relevant evidence | Discussion |
| Generalisability | 21 | Discuss the generalisability (external validity) of the study results | Discussion |
| **Other information** |  |  |  |
| Funding | 22 | Give the source of funding and the role of the funders for the present study and, if applicable, for the original study on which the present article is based | Title page |

